# Digital Cognitive Behavioral Therapy in Anxiety Disorders: A Review of Efficacy and Accessibility

**DOI:** 10.1101/2025.08.08.25333326

**Authors:** Shaunt Papelian

## Abstract

**Background:** Anxiety disorders are among the most prevalent mental health conditions globally, significantly impacting individuals’ quality of life and contributing to the global burden of disease. Cognitive Behavioral Therapy (CBT) is an evidence-based intervention, but traditional in-person therapy faces numerous barriers, including cost, access, stigma, and shortage of providers.

**Objective:** This review explores the growing field of digital CBT (dCBT) for anxiety disorders, summarizing recent findings on its clinical efficacy, accessibility, and future integration into healthcare systems.

**Methods:** A structured search was conducted of studies published from 2017 to 2024 in PubMed, PsycINFO, and Scopus databases, focusing on randomized controlled trials (RCTs), meta-analyses, and large observational studies on dCBT for anxiety.

**Results:** Multiple RCTs and meta-analyses have demonstrated that dCBT is significantly effective in reducing generalized anxiety disorder (GAD), social anxiety disorder (SAD), panic disorder (PD), and specific phobias, often with moderate-to-large effect sizes. Key advantages include scalability, anonymity, and cost-effectiveness. However, dropout rates and patient adherence remain concerns.

**Conclusions:** Digital CBT is a viable alternative or supplement to in-person therapy for anxiety disorders. Further research is needed on personalizing interventions and optimizing engagement.

## 1. Introduction

Anxiety disorders affect over 300 million people globally and are the most prevalent class of mental health conditions. Despite the availability of effective treatments such as Cognitive Behavioral Therapy (CBT), many individuals remain untreated due to barriers like cost, stigma, limited clinician availability, and geographic inaccessibility.

Digital CBT (dCBT), defined as CBT delivered via mobile apps, web platforms, or computer-based programs, has emerged as a scalable and accessible solution. Particularly during the COVID-19 pandemic, the adoption of digital mental health tools increased dramatically. This review explores the clinical effectiveness, accessibility, and limitations of dCBT in the treatment of anxiety disorders.

## 2. Methods

### 2.1. Search Strategy

A systematic search was conducted using PubMed, PsycINFO, and Scopus with the following terms: (“digital CBT” OR “internet-based CBT” OR “online CBT”) AND (“anxiety” OR “generalized anxiety disorder” OR “social anxiety” OR “panic disorder” OR “phobia”). Articles published between January 2017 and June 2024 were considered.

### 2.2. Inclusion Criteria

Eligible studies were peer-reviewed randomized controlled trials (RCTs), meta-analyses, or large-scale cohort studies that involved adult participants over the age of 18. The digital CBT intervention had to be the primary or sole treatment used. The studies were required to report anxiety reduction as a primary outcome, measured using validated anxiety rating scales.

### 2.3. Exclusion Criteria

Studies were excluded if they relied solely on non-CBT frameworks such as mindfulness-only apps, focused on children or adolescents under the age of 18, or used blended therapy approaches without separating dCBT outcomes from other modalities.

## 3. Results

### 3.1. Clinical Efficacy

A 2022 meta-analysis by Carlbring et al. including 3,800 participants reported a pooled Hedge’s g = 0.78 for dCBT in reducing anxiety symptoms, indicating a large effect size. A 2021 randomized controlled trial by Andersson et al. found that dCBT was non-inferior to face-to-face CBT in treating both generalized anxiety disorder and social anxiety disorder.

Evidence suggests that dCBT is effective for a variety of anxiety conditions, including panic disorder and specifiic phobias. The effectiveness is often enhanced when dCBT is supplemented with therapist guidance, even when communication is asynchronous.

### 3.2. Dropout and Adherence

Dropout rates for digital CBT programs range from 25 percent to 50 percent. Therapist involvement has been shown to reduce these rates substantially. Factors contributing to non-adherence include poor user interface design, a lack of personalization, and user fatigue. Digital interventions that incorporate gamifiication features, adaptive content delivery, and social support components show higher completion rates.

### 3.3. Accessibility and Health Equity

Digital CBT has proven especially useful in expanding mental healthcare access to underserved populations, including those living in rural areas and individuals from low-income backgrounds. Several studies have reported successful outcomes even when dCBT programs were offered at no cost. Popular mobile apps such as Sanvello and MindShift have gained traction, particularly among university students and young adults.

Additional features that enhance accessibility include mobile device compatibility, offline access, and availability of materials in multiple languages. Nevertheless, challenges remain in reaching individuals who lack digital literacy or stable internet connections.

## 4. Discussion

The current evidence base strongly supports digital CBT as an effective and practical treatment for anxiety disorders. Although traditional in-person CBT continues to play a critical role, dCBT offers key advantages. These include reduced cost of delivery, greater privacy and convenience for users, and the ability to scale services across large populations without significant increases in infrastructure or staffing.

Despite its benefits, dCBT implementation is not without limitations. High dropout rates can diminish the effectiveness of interventions over time. Furthermore, not all digital programs are developed based on evidence or evaluated through clinical trials. Promising solutions involve the use of hybrid models that combine minimal human guidance with AI-driven personalization.

Future research must examine the role of demographic and psychosocial factors in shaping engagement and efficacy. Studies should also investigate the most effective strategies for integrating dCBT into primary care workflows and health systems.

## 5. Conclusion

Digital CBT represents a compelling and accessible approach to the treatment of anxiety disorders. Its efficacy, particularly when therapist guidance is included, compares favorably with traditional face-to-face therapy. To maximize its potential, stakeholders must focus on improving user engagement, ensuring clinical validity, and reducing disparities in digital access.

If these challenges are addressed, digital CBT may serve as a core component of global mental health strategies, offering scalable and equitable care for individuals affected by anxiety disorders.

## Author Contributions

Shaunt Papelian conceptualized, researched, wrote, and reviewed the manuscript.

## Institutional Review Board Statement

Not applicable.

## Informed Consent Statement

Not applicable.

## Data Availability Statement

No new data were created or analyzed in this study. Data sharing is not applicable.

